# Development and validation of a realistic type III esophageal atresia simulator for the training of pediatric surgeons

**DOI:** 10.1101/2024.01.11.24301151

**Authors:** Javier Arredondo Montero, Blanca Paola Pérez Riveros, Oscar Emilio Bueso Asfura, Nerea Martín-Calvo, Francisco Javier Pueyo, Nicolás López de Aguileta Castaño

## Abstract

**Background:** The technical complexity and limited casuistry of neonatal surgical pathology limit the possibilities of developing the necessary technical competencies by specialists in training. Esophageal atresia constitutes the paradigm of this problem. The use of 3D models for training is a promising line of research, although the literature is limited.

**Methods:** We conceptualized, designed, and produced an anatomically realistic model for the open correction of type III oesophageal atresia. We validated it with two groups of participants (experts and non-experts) through face, construct, and content validity questionnaires.

**Results:** The model was validated by 9 experts and 9 non-experts. The mean procedure time for the experts and non-experts groups was 34.0 and 38.4 minutes respectively. Two non-experts did not complete the procedure at the designed time (45 minutes). Regarding the face validity questionnaire, the mean rating of the model was 3.2 out of 4. Regarding the construct validity, we found statistically significant differences between groups for the equidistance between sutures, 100% correct in the expert group vs. 42.9% correct in the non-expert group (p=0.02), and for the item “Confirms that tracheoesophageal fistula closure is watertight before continuing the procedure”, correctly assessed by 66.7% of the experts vs. by 11.1% of non-experts (p=0.05). Concerning content validity, the mean score was 3.3 out of 4 for the experts and 3.3 out of 4 for the non-experts (p=0.32).

**Conclusions:** The present model is a cost-effective, simple to produce, and validated option for the training of open correction of type III oesophageal atresia. Future studies with larger sample sizes and blinded validators are needed before drawing definitive conclusions.

**Funding:** None

## Introduction

Surgery is a constantly developing and expanding field. The advent of minimally invasive techniques and highly complex procedures (such as robotic surgery or microsurgery) has brought significant improvements for patients in terms of overall health outcomes, hospital stay, and perioperative morbidity, but it has also significantly increased the learning curves of the procedures [1–4]. In pursuit of the best outcome for patients, the global trend has been to centralize these procedures in high-volume centers [5,6]. While this has proven beneficial in multiple pathologies, it has also prevented many of these procedures from being trained and learned by the bulk of surgical specialists. These problems are especially relevant in the field of Pediatric Surgery, where we find additional challenges such as the reduced size of the surgical fields, the tissue fragility, or the intrinsic lability of young patients in terms of homeostasis, which requires procedures to be carried out quickly and efficiently. And all these challenges reach their highest expression in neonatal surgery.

One of the main problems in neonatal surgery is the limited caseload. Although it varies according to pathology, it is common to find a lower number of cases than estimated necessary to acquire basic skills in the management of these pathologies. The absence of a specific guide for trainees that specifies how many procedures of each pathology he/she must attend and perform before the end of his or her training period also contributes to this problem. In addition, the development of minimally invasive techniques in neonatal surgery (such as thoracoscopic correction of esophageal atresia) has led to a tendency for these pathologies to be centralized and to be operated by senior consultants, which limits the training possibilities for residents. The different training models depending on each country also condition this training: there are places like Spain where Pediatric Surgery is a complete surgical specialty with a 5-year training, while in other centers Pediatric Surgery is a fellowship derived from General Surgery [7,8].

Of all neonatal surgical pathologies, esophageal atresia constitutes the paradigm: its high technical complexity, the potential sequelae of surgical iatrogenesis, and the need for a fast and efficient procedure due to the respiratory and hemodynamic lability of the patients justify the need for prior targeted training before performing the procedure in humans. There are few precedents for neonatal thoracic and esophageal atresia models for surgical training [9–15]. Under this premise, we conceptualized, developed, and validated the present model of type III esophageal atresia (esophageal atresia with distal tracheoesophageal fistula).

## Methods

### Conceptualization and preliminary design of the model

For the initial design, anthropometric references of the esophagus, the azygos vein, and the trachea were taken from different bibliographic references [16,17], and a preliminary range of measures and diameters was established. Autodesk Fusion 360 ® (Autodesk, CA., USA) was used to design a first topographic composition of the model and to establish an approximate proportional relationship between the different elements that integrated it. This modeling phase was characterized by iterative adjustments and refinements, ensuring the model’s high fidelity to actual neonatal esophageal conditions.

### Model production methodology

The simulator’s soft base was created with 50g of Eco-Flex 0030 silicone with red dye. 3D printed components, including the bases for the esophagus and trachea and side caps, were produced using PLA (polylactic acid) (Smart Materials 3D, Jaén, Spain) using a Prusa MK3S 3D printer (Prusa Research, Prague, Czech Republic). 3D printed materials were post-processed as required.

The trachea was molded with a Dragon Skin 10 medium silicone (1A:1B, 20g total) with red dye, and the esophagus was molded with an Eco-Flex 0030 platinum silicone with red dye (1A:1B, 5g total). A silicone-based model was chosen instead of a rigid 3D impression to allow a realistic flexible fibro bronchoscopy before the procedure and a realistic closure of the tracheo-oesophageal fistula.

The torso was molded using a Sorta Clear bicomponent silicone with red dye (1A:1B, 150g total), covered by a layer of Eco Flex 0030 platinum silicone with ‘flesh’ pigment and Eco-Flex 35 Fast silicone for a realistic skin texture. The vena azygos was molded using the same silicone mixture as the esophagus but with different mixtures (1A:1B, 10g total).

In relation to the assembly process, the silicone torso was attached to the side covers and it was trimmed to fit correctly. The tracheoesophageal fistula was manually made up ensuring a homogeneous distribution of the silicone sealant. The trachea and the esophagus were inserted into the main base and secured with nylon flanges. The azygos vein was positioned in the desired position by drilling into the silicone and PLA bases and by inserting the silicone tube in the holes. After that, it was filled with water tinted with a drop of methylene blue and subsequently sealed.

### The final version of the model

The present model is constituted by a rectangular surface padded with silicone on which is installed a plastic frame that anchors the essential elements of the model: 1) an anatomically realistic, neonatal-sized trachea, with tracheal rings, a carina, and two main bronchi with a well-delimited *Pars Membranosa* (of 0.8 mm). The internal diameter of the trachea was 8mm and the external diameter was 10.4 mm at the level of the tracheal rings and 9.6 mm at the level of the space between tracheal rings, and 2) an atretic esophagus with a blind proximal pouch, and a distal tracheoesophageal fistula inserted into the trachea with a permeable lumen and conical morphology, simulating real surgical conditions. The esophagus has an outer diameter of 8mm and an inner diameter of 6mm. 3) an azygos vein located over the distal tracheoesophageal fistula, with an internal diameter of 5mm, filled with water with dye (methylene blue).

The model allows regulation of the separation between the two esophageal ends to perform the procedure with different degrees of tension and allows a diagnostic flexible bronchoscopy with a fibrobronchoscope up to 4 mm. Externally, the model is covered by a convex multilayer semi-rigid silicone that simulates the neonatal thorax. On this silicone, it is possible to perform any desired approach (either thoracoscopic or using a variable-size thoracotomy). The simulator’s general dimensions are 146mm x 82mm x 63mm.

The production time for the complete model is 3 hours. The production time for the replacement parts (silicone) is 2 hours. The production cost of the model considering materials, labor, and indirect costs (electricity consumption) is 52.58 € for the complete model and 35.86 € for the model without the molds. The cost of each spare part (azygos vein, esophagus, and trachea) is 17.05 €.

### Validation

A validation protocol was designed and implemented with two groups of validators: 1) experts (group 1) and 2) non-experts (group 2). Experts consisted of consultant Pediatric Surgeons who had performed the procedure at least once. The non-expert group was considered to have basic surgical skills, but no specific training in neonatal surgical pathology or in the surgical management of oesophageal atresia. Therefore, the non-expert group consisted of second-to-fifth year General or Pediatric Surgery residents.

The validators (OEB and BPR) were specifically instructed in the pathology, the surgical technique, the validation questionnaires, and the elements to be assessed during the performance of the procedure. All validations were supervised by them.

Both experts and non-experts had never had previous contact with the model. For the validation process, specific questionnaires and checklists were designed for construct, content, and face validity.

The validation process had the following structure: 1) first, an illustrative video was reproduced, showing with detail the surgical procedure to be performed with a demonstration of the use of the model; 2) validation was initiated under direct visual supervision of the validators and with continuous recording of the surgical field in case a re-evaluation of the validations was required.

We would like to express our sincere gratitude to the following people for their altruistic and voluntary participation in the validation of this simulator. Without their feedback and willingness, this project would never have been possible:

“the names of the participants have been blinded for peer review, but will appear in the final version of the manuscript”

## Results

The model was validated by 9 experts (group 1) and 9 non-experts (group 2). The mean procedure time for the experts and non-experts group was 34 (sd=5.8) and 38.4 (sd=6) minutes respectively. Two non-experts did not complete the procedure on the proposed time limit, which was 45 minutes.

### Construct validity

All the experts (n=9) and non-experts (n=9) responded to the construct validity questionnaire. We found statistically significant differences for the equidistance between sutures, which was 100% correct in the expert group vs. 42.9% correct in the non-expert group (p=0.02) and for the item “Confirms that tracheoesophageal fistula closure is watertight before continuing the procedure”, which was correctly assessed by 66.7% of the experts vs. by 11.1% of non-experts (p=0.05). Table 2 shows the comparison between groups for construct validity items.

### Face validity

All the experts (n=9) responded to the face validity questionnaire. All items were rated on a scale from 1 (worst) to 4 (best). The best-rated aspect was “this model allows the acquisition of surgical skills transferable to the real surgical field”, with a score of 3.7. The worst-rated item was “the azygos vein resembles that of a neonate”, with a score of 2.4. The mean rating of the model was 3.2 out of 4 (sd=0.27) Table 1 shows the mean score attributed to each of the items in the questionnaire.

### Content validity

All the experts (n=9) and non-experts (n=9) responded to the content validity questionnaire. All items were rated on a scale from 1 (worst) to 4 (best). In the experts group the best-rated aspect was “this model allows the user understanding the procedure technique”, with a mean score of 3.8 and the worst-rated aspect was “this model allows the user to auto-evaluate its capacity to perform this procedure” with a mean score of 2.9. In the non-experts group the best-rated aspect was “this model allows to learn different surgical techniques” with a mean score of 3.8 and the worst-rated aspect was “this model allows the user to auto-evaluate its capacity to perform this procedure” with a mean score of 3. The mean rating of the model was 3.3 out of 4 for the experts (sd=0.28) and 3.4 out of 4 for the non-experts (sd=0.22). Table 3 shows the mean score attributed to each of the items in the questionnaire.

## Discussion

Through the present work, our pediatric surgical simulation development group (SIMUPED ®) conceptualized, designed, produced, and validated an anatomically realistic model of type III oesophageal atresia for the training of pediatric surgeons in training. The limited casuistry of this pathology, the lability of the patients who suffer from it, and the high technical complexity of the corrective procedure justify the development of training models to acquire the necessary skills and competencies before being able to perform the procedure on humans. The model we have developed stands out for being anatomically realistic, having a low production cost (both in terms of time and materials), and allowing the performance of bronchoscopy.

In this validation study, no differences were found in most items between experts and non-experts for the construct validity items, except for the equidistance between sutures and the confirmation of the closure of the fistula. We attribute these findings to the fact that this procedure is highly technical and specialized and required extensive pre-procedural guidance and instructions. We believe this pre-procedural guidance may in part have hampered our results. It is pertinent to note that 2 non-experts did not complete the procedure in time, which also may have hampered our results. If we considered a hypothetical completion time of 46 minutes for these two non-experts (one minute more than the proposed limit), the difference in time between the two groups would have reached marginal significance (p=0.06). The face validity obtained an overall positive evaluation, with a score of 3.2 out of 4. The lowest-rated item, the azygos vein anatomy, corresponds to the most difficult structure to replicate due to the technical limitation of generating an anatomically realistic hollow tubular structure filled with liquid. However, we consider that its consistency and the presence of liquid allowed to train the dissection and ligation technique. Finally, concerning content validity, the overall assessment of the model was positive, with an average of 3.4 out of 4 in the group of experts and 3.4 out of 4 in the group of non-experts. The fact that the most highly rated items by the experts was that the model allowed understanding of the technique is very positive in terms of the transferability of the model.

The validation study faced some difficulties worth mentioning. 1) First, the definition of expert and non-experts was complicated: the non-experts must have some basic surgical skills to be able to carry out the procedure but they had to be sufficiently different from the experts so that the model could distinguish them from the non-experts in the construct validity questionnaire. It may be that the groups were not sufficiently different or that the questions included in the construct validity questionnaire did not accurately reflect the expert character. However, we found significant differences in items of great relevance (such as the equidistance between points) that we believe serve to support the constructive validity of the model. 2) Second, due to the scarce number of experts in the hospitals in our area, we were unable to recruit more participants.

To our knowledge, 7 models of oesophageal atresia have been previously published, all of them between 2014 and 2023. A summary of the main characteristics of the models published to date is shown in Table 4. Except for the first one, developed by Barsness et al. and using bovine fetal tissue, all subsequent ones have been synthetic. Regarding the surgical technique aimed, the model by Neville et al. was designed expressly for open surgery, and all the others were designed for thoracoscopic procedures. To our knowledge, the one presented in this work is the first model that allows the performance of a realistic flexible bronchoscopy. This involves multiple aspects of interest, such as specific training in neonatal bronchoscopy or endoscopic treatment of tracheoesophageal fistula. Variants of this model aimed at training and developing these competencies are of interest in the future. One of the strengths of our model is the use of silicone to replicate the trachea because it results more realistic than 3D-printed ones. Another highlight of our model is that since the tracheoesophageal fistula is made of silicone and placed manually, all types of oesophageal atresia could be simulated by introducing slight variations in the model. This is of great value if one wanted to train the technique blind to the type of atresia, which is the situation in which most surgeons find themselves before starting an operation of this type. In addition to having a lower cost, an important contribution of our model compared to the existing ones is the inclusion of the azygos vein. We also believe that the possibility of regulating the tension of the oesophageal cords is a notable contribution to the previous models since it allows to vary the difficulty of the practice.

Our model is oriented to the open correction of the atresia: although the model allows the procedure to be performed thoracoscopically, we believe that it is essential that all pediatric surgeons receive adequate training in the open correction of oesophageal atresia, and this is, to the best of our knowledge, the first model oriented along these lines.

Lastly, although we considered the possibility of testing the anastomosis for leakage after completion of the procedure, the laboratory tests we performed showed leakage through the needle insertion and passage points, and therefore it was not feasible. We attributed this to the intrinsic characteristics of the silicone. This should be considered a limitation of our model and a potential area for improvement in subsequent designs.

Regarding the cost of our model, it should be considered that the price was estimated for individual hand-made production. We believe that the production process of our model is industrialisable, which would result in greater uniformity and lower labor costs. The price would become much more competitive since the bulk of the price we report here is labor.

About the validation studies of the previously published models, we found great heterogeneity. Although the reported results are in general good, we identified several discrepancies in the definition of the groups of experts and non-experts, in the sample sizes, and the validation methodology used. We consider the inclusion of a group of non-experts to be a strength of our work, given that in many of the previous articles, there was only a group of experts. We also believe that the design of construct validity questions is complex and would benefit in the future from collaborative groups for their conception (e.g. through the Delphi method)

This study is not exempt from limitations. First, the limited sample size may have hampered the obtention of more statistically significant differences between groups. Second, the validator was not blinded to the type of participant, which may have biased his/her responses. Third, the inclusion of an additional external validation would have been useful to assess the reproducibility of the construct validity questionnaire. On the other hand, the main strength of this study is the detailed reporting of the methodology and measurements of the model, which facilitates its reproducibility.

In conclusion, the present model is a low-cost, simple-to-produce, and validated option for the training of open correction of type III oesophageal atresia. Future studies with larger sample sizes and blinded validators are needed to draw definitive conclusions.

**Figure 1.**
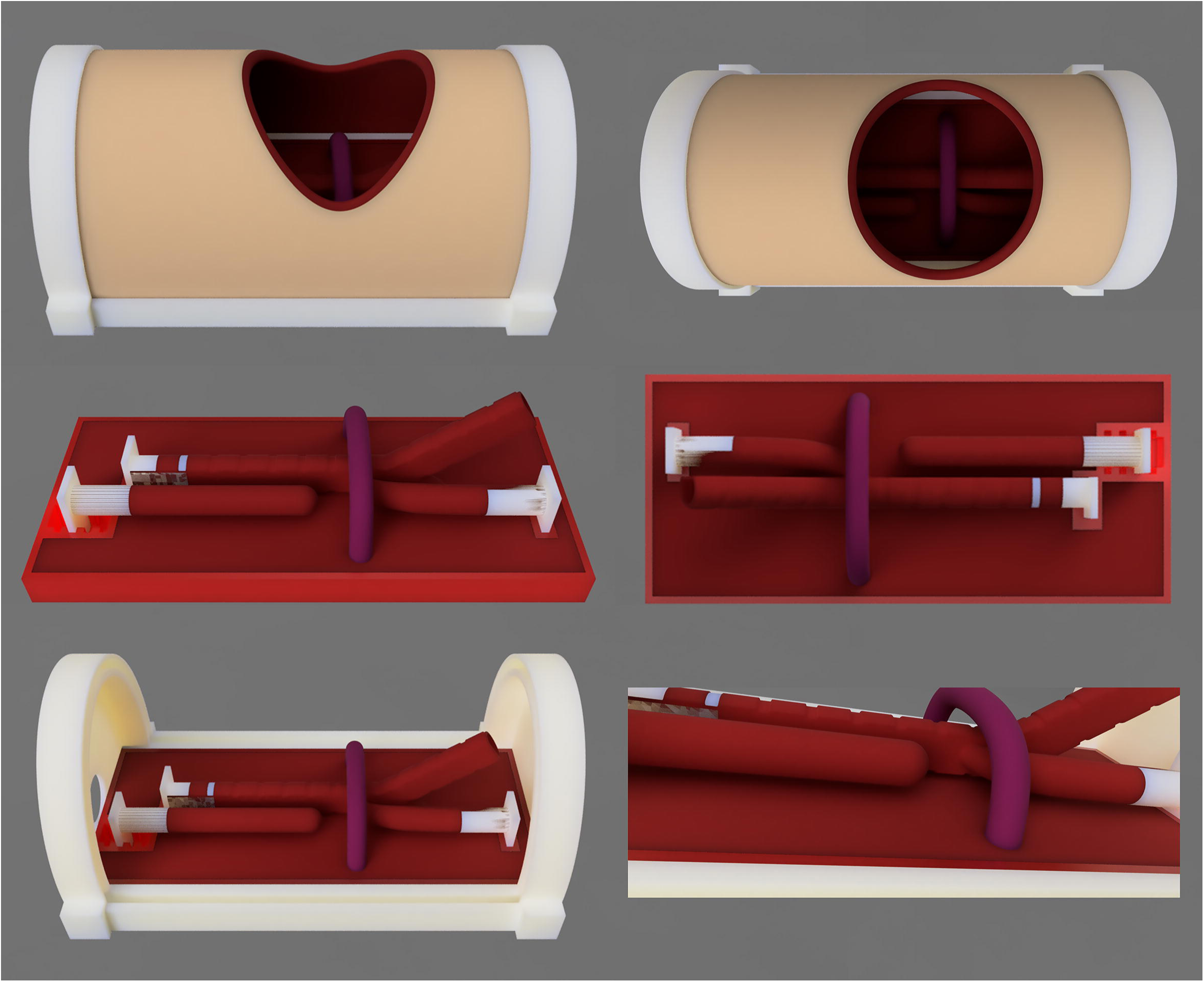
Images corresponding to the design phase of the model, made in Autodesk Fusion 360 ® (Autodesk, CA., USA).

**Figure 2.**
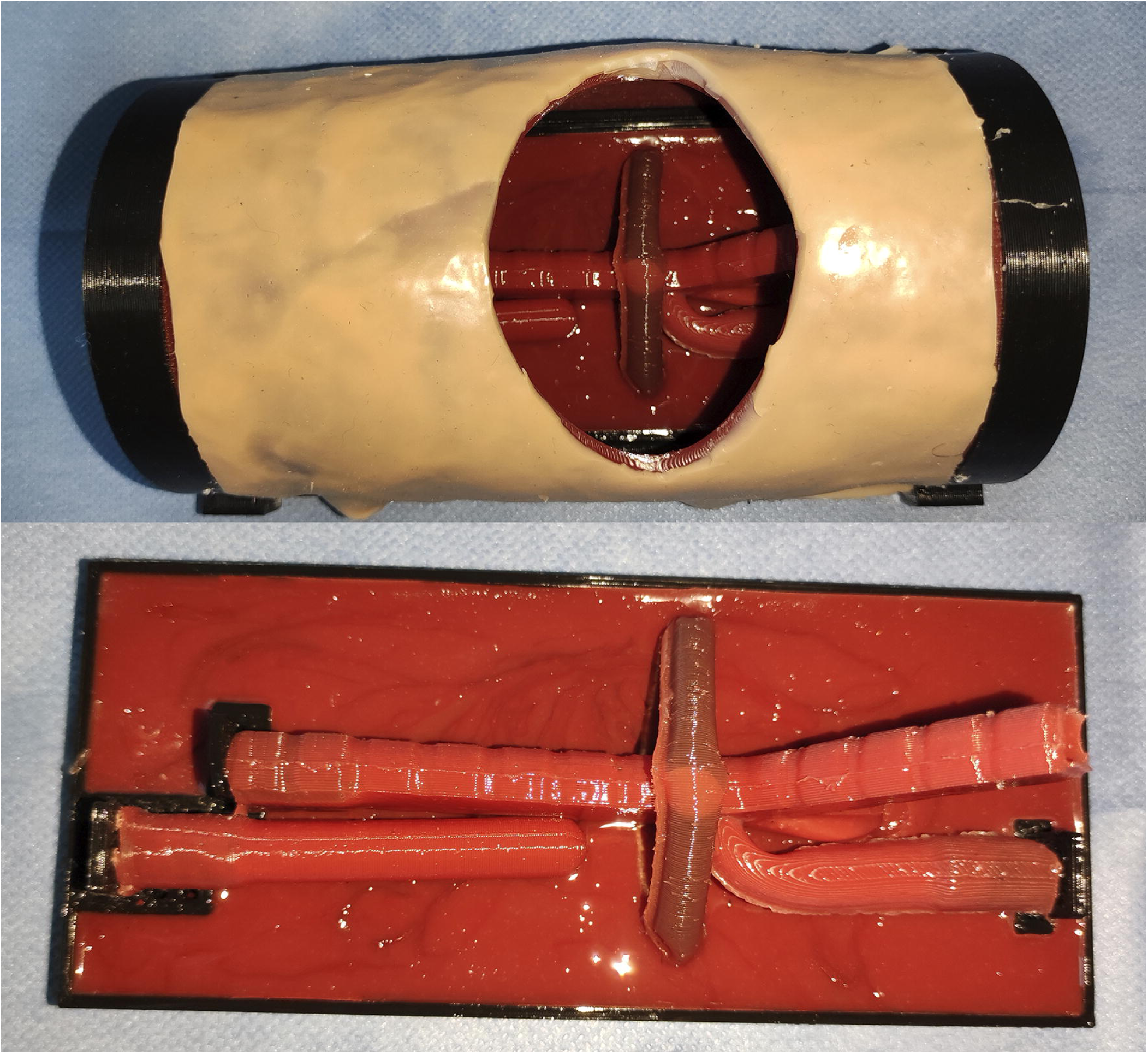
**Above**: Appearance of the final model with the thoracic coverage. **Below**: Appearance of the model without the thoracic coverage.

**Figure 3.**
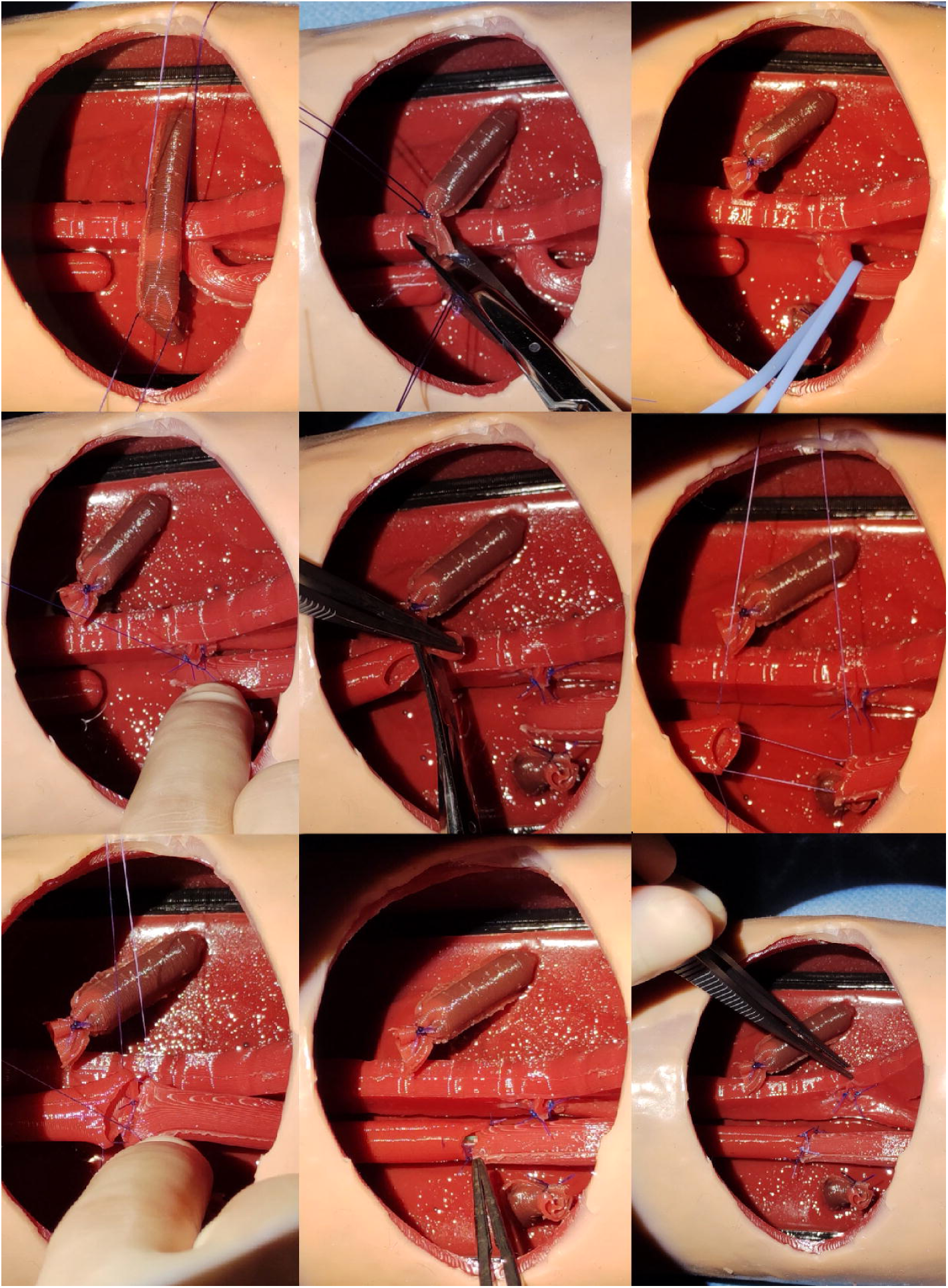
Representative images of the performance of the oesophageal atresia open correction on the model. **Above left, center**: azygos vein reference and ligation; **Above right, center left**: tracheo-esophageal fistula reference and ligation. **Center, center**: opening of the upper esophageal pouch; **Center, right**: lateral sutures of the oesophageal anastomosis; **Bottom left, center**: oesophageal anastomosis. **Bottom, right**: final result.

## Data Availability

The data used to carry out this study are available upon request from the review authors.

## CRediT authorship contribution statement

**JAM:** Original idea; literature search; model conceptualization and design; study design; data curation and extraction; project administration; resources; writing – original draft; writing – review and editing.

**NLC:** Model conceptualization and design; model production, project administration; resources; writing – original draft; writing – review and editing.

**NMC:** Study design; data curation and extraction; formal analysis; project administration; resources; writing – original draft; writing – review and editing.

**BPR, OBA:** Model production and validation; analysis; resources; writing - original draft; writing - review and editing.

## ETHICAL APPROVAL

This project was approved by the Research Ethics Committee of the University of Navarra on 13.4.2023 under code 2023.059.

## STATEMENT OF AVAILABILITY OF THE DATA USED

The data used to carry out this study are available upon request from the review authors.

## ACKNOWLEDGEMENTS

– Alvira Reyes Delgado
– Paolo Bragagnini
– Ainara González Esdera
– Paulina Vargova
– Yurema González Ruiz
– Marina González Herrera
– Mercedes Ruiz de Temiño Bravo
– Carolina Corona Bellostas
– Rafael Fernández Atuan
– Raquel Ros Briones
– Lucas Sabattel
– Nuria Blanco
– Sara Hernández-Martín
– Elena Calleja Aguayo
– Carlos Bardají Pascual
– Helena Linero González
– Iker Rodríguez Laguna
– Cristian Fernández Romance
– Carmen María Gálvez Estévez
– Marina Román Moleón

