## Supplementary material for "Development and validation of a realistic type III esophageal atresia simulator for the training of pediatric surgeons": Table 1

| **Item** | **Mean score (experts)**  **N=9** |
| --- | --- |
| The esophageal diameter of the model resembles that of type III esophageal atresia (esophageal atresia with distal tracheoesophageal fistula). | 3.4 |
| The esophageal thickness of the model resembles that of type III esophageal atresia (esophageal atresia with distal tracheoesophageal fistula). | 3.3 |
| The trachea resembles that of a neonate. | 2.9 |
| The azygos vein resembles that of a neonate. | 2.4 |
| The elements of the model are adequately represented and laid out | 3.1 |
| The layers of the model realistically simulate a real esophagus. | 3.2 |
| Visually, the model material resembles a real esophagus. | 3 |
| The feel of the model (wet and slippery) is reminiscent of a real esophagus. | 3.1 |
| The texture and consistency of the model when the suture needle passes through it is similar to that of a real esophagus. | 3.3 |
| The sensation of suturing the ends of the model is similar to that of a real esophagus. | 3.1 |
| The model reproduces the surgical dimensions of a neonatal thoracic field. | 3.4 |
| The spatial positioning of the model resembles that of a surgical field. | 3.1 |
| The model allows a mobility of the structures similar to reality. | 3 |
| In general, the execution of the surgical technique for the correction of a type III esophageal atresia on the model resembles reality | 3.3 |
| This model is USEFUL for LEARNING the surgical technique for the correction of type III esophageal atresia. | 3.4 |
| This model allows a realistic simulation of an end-to-end esophageal anastomosis (moderate tension). | 3.3 |
| This model allows the acquisition of surgical skills transferable to the real surgical field. | 3.7 |
| This model realistically reproduces the level of difficulty of the procedure. | 3.1 |
| **MEAN SCORE** | **3.2** |

**Table 1. Face validity questionnaire.**

Each item was evaluated on a 4 point scale (best rating 4, worst rating 1)
