## Supplementary material for "Development and validation of a realistic type III esophageal atresia simulator for the training of pediatric surgeons": Table 2

| **Item** | **Experts (n=9)** | **Non-experts (n=9)** | **p-value** |
| --- | --- | --- | --- |
| Decides to ligate the azygos vein before dissecting the esophagus (% yes) | 100 | 100 | . |
| Ligates the azygos vein without tearing it (% yes) | 77.8 | 77.8 | 0.99 |
| Correctly identifies both esophageal pouches and tracheoesophageal fistula (% Yes) | 88.9 | 88.9 | 0.99 |
| Checks by extrinsic collapse that the tracheoesophageal fistula has been correctly identified (% Yes) | 66.7 | 22.2 | 0.15 |
| Closes tracheoesophageal fistula progressively (cut-point-cut) (% Yes) | 77.8 | 88.9 | 0.99 |
| Resects only the essential amount of esophagus at the level of the tracheoesophageal fistula (% Yes) | 100 | 88.9 | 0.99 |
| **Confirms that tracheoesophageal fistula closure is watertight before continuing the procedure (% Yes)** | **66.7** | **11.1** | **0.05** |
| Ensures that there are no additional tracheoesophageal fistulas (% Yes) | 77.8 | 55.6 | 0.62 |
| Adequately dissects both esophageal ends (% Yes) | 100 | 100 | . |
| Checks the distance between esophageal ends and the viability of the anastomosis before performing the anastomosis (% Yes) | 88.9 | 55.6 | 0.29 |
| Starts with side sutures and references them (% Yes) | 100 | 100 | . |
| Then performs the posterior face anastomosis (% Yes). | 100 | 88.9 | 0.99 |
| Places a transanastomotic tube before suturing the anterior face (% Yes) | 77.8 | 55.6 | 0.62 |
| In general, lowers the knots with a controlled and homogeneous tension and accompanies them by the dominant finger (% Yes). | 100 | 88.9 | 0.99 |
| In general, sutures are full thickness (% Yes) | 100 | 87.5 | 0.47 |
| **In general, sutures are equidistant from each other (% Yes)** | **100** | **42.9** | **0.02** |
| In general, it adequately lowers knots (“surgeon´s knot”) (% Yes) | 100 | 100 | . |
| Completes the procedure (% Yes) | 100 | 77 | 0.47 |
| Mean number of sutures | 3.1 | 3.7 | 0.35 |
| Mean procedure time (minutes) | 34 | 38.4 | 0.19 |

**Table 2. Construct validity evaluation.**
