## Supplementary material for "Development and validation of a realistic type III esophageal atresia simulator for the training of pediatric surgeons": Table 3

| **Item** | **Mean score (experts)**  **N=9** | **Mean score (non-experts)**  **N=9** | **p-value** |
| --- | --- | --- | --- |
| This model helps the user understanding the technique behind the procedure. | 3.8 | 3.4 | 0.54 |
| This model helps the user understanding the spatial arrangement of the thorax and mediastinum in a neonate. | 3.4 | 3.3 | 0.64 |
| This model helps the user to learn how to handle the esophagus and neonatal mediastinal structures in a surgical context. | 3 | 3.2 | 0.33 |
| This model helps the user understanding how esophageal and tracheal tissue responds to being handled in surgery. | 3 | 3.3 | 0.12 |
| This model helps the user understanding how esophageal tissue responds to an end-to-end anastomosis under moderate tension. | 3.2 | 3.3 | 0.76 |
| This model allows to LEARN different surgical techniques | 3.2 | 3.8 | 0.11 |
| This model allows TRAINING of different surgical techniques | 3.6 | 3.6 | 0.78 |
| This model allows to EVALUATE the user's surgical technique. | 3.2 | 3.3 | 0.76 |
| This model is used to measure the user's ability to perform corrective surgery for esophageal atresia type III (esophageal atresia with distal tracheoesophageal fistula). | 2.9 | 3 | 0.65 |
| This model helps the user to be better prepared when performing corrective surgery for esophageal atresia type III (esophageal atresia with distal tracheoesophageal fistula) in a neonate for the first time. | 3.6 | 3.6 | 0.8 |
| This model serves to increase the user's confidence before performing corrective surgery for type III esophageal atresia (esophageal atresia with distal tracheoesophageal fistula) in a neonate. | 3.4 | 3.6 | 0.65 |
| **MEAN SCORE** | **3.3** | **3.4** |  |

**Table 3. Content validity questionnaire.**

Each item was evaluated on a 4 point scale (best rating 4, worst rating 1)
