## Supplementary material for "Development and validation of a realistic type III esophageal atresia simulator for the training of pediatric surgeons": Table 4

| **Author** | **Country** | **Model type** | **Technique** | **Validators** | **Validation items** | **Validation results** | **Model cost** | **Model measures** |
| --- | --- | --- | --- | --- | --- | --- | --- | --- |
| Barsness et al. (2014) | USA | Mixed (synthetic + bovine fetal tissue) | Thoracoscopy | -Non-experts: self-reported “novice” PS (n=12)  -Experts: self-reported “experienced” PS (n=8) | -24 item content validity survey  -OSATS – two independent PS | -High overall simulator ratings  -OSATS: High interitem consistency  -OSATS: High interrater agreement | NR | NR |
| Harada et al. (2016)* | Japan | Synthetic | Thoracoscopy | PS (n=4) | -Optical and electromagnetic tracking  -Task completion time  -Content validity survey | -High overall simulator ratings  -High tracking ratio (99.8%) | NR | NR |
| Maricic et al. (2016) | Argentina | Synthetic | Thoracoscopy | -Experts (PS with >30 TEF/EA repairs) (n=7)  -Intermediates (PS with 5 to 29 TEF/EA repairs) (n=10)  -Beginners (PS with <5 TEF/EA repairs) (n=22)  Total: n=39 | -Likert-type scale  -Performance checklist | -High overall simulator ratings  -Significant differences in time and number of errors between Experts/intermediates and beginners | Low cost  (not specified) | NR |
| Deie et al. (2016)* | Japan | Synthetic | Thoracoscopy | -Experts (PS with ≥ 3 TEF/EA thoracoscopic repairs) (n=6)  -Non-experts (PS with < 3 TEF/EA thoracoscopic repairs) (n=34)  -Validators divided also between ESSQ qualified (n=15) vs ESSQ non-qualified (n=25) | -Questionnaire (from Barsness et al.)  -Video-based endoscopic surgical skill assessment:  29-item checklist  Error assessment sheet | -Experts significantly superior to non-experts | NR | NR |
| Neville et al. (2022) | UK | Synthetic | Open repair | -Experts (n=12)  -Non-experts (n=28) | -Construct validity  -Content validity  -Face validty | -High overall simulator ratings  -Experts significantly superior to non-experts | Structure: 100￡  Replaceable parts:  20￡ per use | NR |
| Zahradniková et al. (2023) | Slovakia | Synthetic | Thoracoscopy | -Experts (experienced surgeons): n=7  -Intermediates: Pediatric Surgery trainees (n=4)  -Novices: Medical students (n=7) | -Face validity  -Content validity  -Overall impression  -OSATS  -Quality of the anastomosis | -High overall simulator ratings | NR | NR |

**Table 4. Summary of the esophageal atresia training models published to date**

UK: United Kingdom; USA: United States of America; PS = Pediatric Surgeons; OSATS: Objective Structured Assessments for Technical Skills; NR: not reported; TEF/EA: Esophageal atresia with tracheoesophageal fistula; ESSQ: Endoscopic Surgical Skill qualification; ID: Inner diameter; OD: Outer diameter

*: same working group
